# Assessment tools and incidence of hospital-associated disability: a rapid systematic review

**DOI:** 10.1101/2022.09.22.22279726

**Authors:** Katia Giacomino, Roger Hilfiker, David Beckwée, Jan Taeymans, Karl Martin Sattelmayer

**Affiliations:** School of Health Sciences, HES-SO Valais-Wallis, Leukerbad, Valais-Wallis, 3954, Switzerland; Rehabilitation Research (RERE) Research Group, Department of Physiotherapy, Human Physiology and Anatomy, Faculty of Physical Education and Physiotherapy, Vrije Universiteit Brussel, Brussels, Belgium; Division of Physiotherapy, Department of Health Professions, University of Applied Sciences Bern, Bern, Switzerland; Faculty of Physical Education and Physiotherapy, Vrije Universiteit Brussel, Belgium

**Keywords:** Functional decline, hospitalization, activities of daily living, older patients

## Abstract

**Background:** During hospitalization older people have a high risk of developing functional impairments unrelated to the reasons for their admission. This is termed hospital-associated disability. This systematic review aimed to assess the incidence of hospital-associated disability among older patients admitted to acute care, to identify the tools used to assess activities of daily living in these patients, and evaluate which functional task is most sensitive for detecting changes in disability among older hospitalized patients.

**Methods:** A rapid systematic review was performed according to the recommendations of the Cochrane Rapid Reviews Methods Group and the PRISMA statement. A literature search was performed in Medline (via Ovid), EMBASE, and Cochrane Central Register of Controlled Trials databases on 26 August 2021. Inclusion criteria: older people, assessment of activities of daily living at baseline and discharge. Exclusion criterion: diseases affecting functional decline.

**Results:** Eleven studies were included in the final review. Incidence of hospital-associated disability (overall score) was 37% (95% CI 0.31–0.42). Insufficient data prevented meta-analysis of the individual items. The most sensitive measure for detecting changes in disability was the overall score of assessment of activities of daily living.

**Conclusions:** Incidence of hospital-associated disability in older patients might be overestimated, due to the combination of disease-related disability and hospital-associated disability. The tools used to assess these patients presented some limitations. These results should be interpreted with caution, as a limited number of studies reported adequate information to assess the incidence of hospital-associated disability. Risk of bias in the included studies raised some concerns.

## Introduction

Functional decline in older adults during hospitalization increases the risk of a longer hospital stay [1], a nursing home placement [2], and increased mortality [3]. The main goal in older adult care is therefore to maintain function [4] and the ability to perform activities of daily living (ADL) [1, 5].

During hospitalization older adults are at risk of developing functional decline unrelated to the condition for which they were admitted [6]. The loss of independence in at least one activity of daily living is referred to as hospital-associated disability (HAD) [7]. Furthermore, HAD refers to disability acquired during hospitalization or the worsening of a pre-existing disability due to hospitalization [7].

Previous studies have highlighted methodological issues in assessing functional decline [8, 9]. However, there is currently no consensus on which tool should be used to assess functional decline in these patients, which ADL tasks should be included, how the assessment should be performed (self-reported or performance-based), and what time-frame should be considered [8]. Covinsky and others [7] recommend asking patients on admission about their ADL functioning before the onset of acute illness.

A previous study highlighted the magnitude of the HAD problem, and reported that the overall prevalence of HAD among older adults admitted to an acute care hospital is 30% [9]. To the best of our knowledge, the incidence of HAD has not been studied in a systematic review. The aim of this study was to perform a rapid systematic review to: (i) assess the incidence of HAD, (ii) to identify all tools or functional tasks used to assess ADL in hospitalized older patients, and (iii) to evaluate which ADL functional task or set of tasks is the most sensitive to detect changes in disability.

## Methods

### Study design

A rapid systematic review was performed according to the recommendations of the Cochrane Rapid Reviews Methods Group [10]. Reporting was conducted in accordance with the PRISMA statement [11].

### Search strategy and selection criteria

A literature search was performed in Medline (via Ovid), EMBASE, and Cochrane Central Register of Controlled Trials (CENTRAL) databases on 26th August 2021. The search strategy comprised five search terms related to: (i) study setting (i.e. hospital), (ii) observed disability in ADL, (iii) incidence and prevalence, sensitivity to change and responsiveness, (iv) population identification (i.e. older adults), and (v) articles that cover the aspect of disability acquired in hospitals. The search terms, combined with the Boolean operator “AND”, were applied to titles and abstracts, and MeSH terms were added when available and relevant. The full search strategy is shown in Appendix 1.

The review included prospective and retrospective cohort studies. The control group of randomized controlled trials (RCTs) was eligible when performing a usual or a sham intervention. Inclusion criteria were: studies investigating a general older population (≥ 65 years) who were admitted to hospital for an acute disorder; studies had to assess the individual items of the ADL measurement tool before hospitalization (retrospectively or prospectively) and at the end of hospitalization or after hospital discharge.

Exclusion criteria were: studies investigating a specific condition that could have an effect on functional decline (e.g. stroke, brain injury, heart failure, COVID-19, and acute respiratory failure); and studies that primarily examined a population with cognitive impairment.

One reviewer (KG) independently screened all the records based on titles and abstracts (phase 1) and the full-text of the eligible studies (phase 2). A second reviewer (KMS) screened 20% of the same records. If Cohen’s kappa coefficient of agreement was >0.80 for both phases, only 20% of the studies were planned to be screened independently by two reviewers. Disagreements were resolved through discussion.

### Data extraction

Study characteristics (authors, country, study sample size, population age, type of ward, proportion of women, proportion living alone, proportion living in a nursing home, Acute Physiology and Chronic Health Evaluation (APACHE II) score, Charlson Comorbidity Index, number of days of hospitalization, any scale of mental status) were extracted by one reviewer (KG), while incidence data were extracted by three reviewers (RH, KMS, KG) at the same time. Disagreements were resolved through discussion between all three reviewers. The following information was extracted: item type, response options, criteria for the response options, baseline, and discharge assessment (i.e. who performed it and how), baseline and discharge prevalence of ADL dependency, operationalization, and definition of HAD and incidence per item and for the overall score.

### Methodological quality assessment

Methodological quality assessment was performed using the critical appraisal checklist for Studies Reporting Prevalence Data [12] from the Joanna Briggs Institute (JBI). The JBI checklist was completed independently by two reviewers (KMS, KG). Differences in rating were discussed and resolved.

A further five questions were selected from the JBI checklist that were considered to assess risk of bias (internal validity, i.e. questions 2, 5, 6, 7 and 9). Risk of bias was assessed as low if all five questions were rated ‘yes’, some concern if one of the questions was rated ‘unclear’ and high if one of the questions was rated ‘no’.

### Incidence of hospital-associated disability

The incidence of HAD (total score) was pooled using the statistical software R [13] and its package meta [14]. A random effects model was applied based on an inverse variance model with a logit transformation.

Heterogeneity was assessed by the I^2^, which is the proportion of total variability due to between-study heterogeneity [15], to estimate inter-study variability. Tau was estimated with the DerSimonian-Laird estimator [14]. Miglivaca et al. recommended avoiding the use of an arbitrary cut-off for the I^2^ statistics, as it may not be discriminative in incidence studies [16]. Therefore, heterogeneity was explained by discussing the level of dependency at baseline and the method of assessment.

The data required to calculate the incidence of HAD are the number of patients who are dependent and independent at baseline and the evolution of these groups at discharge. If these values were given, the following formula was used to calculate the incidence of the individual ADL task or set of tasks (total score):

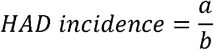

where, a = number of newly dependent patients (requiring the help of someone else), and b = number of patients at risk of developing or increasing dependency.

For example, in the study by Sager et al. [17], 51 patients were newly dependent at discharge according to the overall score and 188 patients had the potential to decline in one of the ADL tasks.

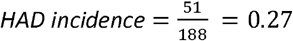

If these values were not reported, an unbiased estimate of the incidence of HAD in that study could not be calculated. In this case, an estimate of the incidence was made by subtracting the percentage of dependency at discharge minus the percentage of dependency at baseline.

### Tools used for assessment of activities of daily living in hospitalized older patients

All ADL tools or sets of tasks used in the assessment of hospitalized older patients were reported narratively.

### Evaluation of which measure is most sensitive to detect changes in disability

The most sensitive task or set of tasks to detect changes in disability were ranked as follows: (i) per study all ADL tasks were ranked from the highest proportion of disability (= 1) to the lowest (=7); (ii) the mean of the ranking was calculated per item or set of scores.

If data were available only in a chart they were extracted using the software WebPlotDigitizer [18]. Due to time constraints, the authors of the included studies were not contacted in case of imprecision in the text.

### Differences from protocol

It was decided not to integrate the grading system of the GRADE Working Group as predefined the evaluation of the body of evidence due to the lack of a guideline for incidence studies.

## Results

A total of 2,519 records were identified (Medline (via Ovid) 740 records, EMBASE 1,557, CENTRAL 222). After removing 743 duplicates, titles and abstracts of 1,776 articles were screened and 1,431 were excluded. The full text of 345 studies was screened, and a final total of 11 studies were included for further analysis [17, 19-28]. The reasons for exclusion are shown in the study flow diagram in Appendix 2.

Cohen’s kappa coefficient was > 0.8 in both screening phases (i.e. titles/abstracts and full texts); therefore only 20% of studies were screened by two reviewers independently, as described above.

### Study characteristics

The population characteristics of the included studies are shown in Table 1. Seven studies were performed in the USA, one in Europe, and three in other countries. Sample size ranged from 36 to 2,877 participants and the age of study participants ranged from 76 to 87 years. The proportion of women in the study samples under investigation ranged from 31% to 66%. Included studies were conducted in the medicine ward (five studies), general hospital (five studies), and one in the geriatric unit and usual unit which refers to a conventional care unit.

**Table 1.** Study characteristics

| Author | Country | Sample size, *n* | Age, years, mean (SD) or (range) | Type of ward | Proportion of women, % | % Living alone | % Living in nursing home | APACHE II score mean (SD) or % of patients per category | Charlson Comorbidity Index mean (SD) or % of patients per category | Mean number of days of hospitalization (SD) or (range) | Mental status mean (SD), tool) or % of patients with cognitive impairment |
| --- | --- | --- | --- | --- | --- | --- | --- | --- | --- | --- | --- |
| Covinsky 2000 [28] | USA | 2877 | 80.5 | Medicine | 64% | 52% | 5.1% | NR | NR | NR | 1.4 (1.3), SPMSQ |
| Covinsky 2003 [27] | USA | 2293 | 79.5 | Medicine | 63.6% | 35.2% | 4.9% | 0–2: 32.3%,<br>3–5 37.1%,<br>>6: 30.8% | 0: 19.8%,<br>1–2: 47.0%,<br>3–4: 22.1%,<br>>5: 11.1% | 6.3 | NR |
| Dharmarajan 2020 [[26] | USA | 515 | 82.7 (5.6) | Hospital | 65.6% | 46.6% | 0% | NR | NR | NR | 17.9%<sup>b</sup>, MMSE |
| Hansen 1999 [25] | USA | 73 | 80.4 (7) | Hospital | 66% | 45% | 0% | NR | NR | 7.5 (5.0) | 2.1%<sup>b</sup>, MMSE |
| Hirsch 1990 [24] | USA | 71 | 84 (75–95) | Medicine | 59% | 7.0% | 5.6% | NR | NR | 10.1 (2–49) | 30.8%<sup>c</sup>; last item of CNA |
| Inouye 1993 [23] | USA | 188 | 78.4 (5.8) | Medicine | 59% | 45% | 4% | 13.9 (3.6) | 8.5 (2.8)<sup>a</sup> | 7<sup>d</sup> (2–51) | 23.5 (5.4), MMSE |
| Martinez-Velilla 2021 [22] | Spain | 149 | 87.1 (5.2) | Hospital | 59% | NR | NR | NR | NR | 8<sup>d</sup> | NR |
| Mudge 2010 [21] | Australia | 615 | 80.4 (7.5) | Medicine | 59% | NR | 11% | NR | NR | 7 (5–13) | 10%, history of dementia |
| Park 2021 [20] | Republic of Korea | RB: 45<br>PF: 36<br>MMF: 37<br>SF: 58 | RB: 77 (73–82)<br>PF: 80 (74–84)<br>MMF: 81 (77–86)<br>SF: 81 (75–84) | Hospital | RB: 31.1%<br>PF: 38.9%<br>MMF: 46.0%<br>SF: 39.7% | NR | RB: 0%<br>PF: 2.8%<br>MMF: 8.1%<br>SF: 53.5% | NR | NR | NR | RB: 82.2%<br>PF: 58.3%<br>MMF: 70.3%<br>SF: 82.8% |
| Sager 1996 [17] | USA | 1279 | 79 (6.3) | Hospital | 62% | 37% | 0% | NR | NR | 8.6 (6.8) | 17 (4.0), MMSE |
| Zelada 2009 [19] | Peru | GU: 68<br>UU: 75 | GU: 79.6 (6.8)<br>UU: 76.1 (7.2) | Geriatric, usual unit | GU: 61.8%<br>UU: 56% | NR | NR | GU: 9 (2.87)<br>UU: 8.4 (3.11) | GU: 3.6 (1.98)<br>UU: 3.1 (1.6) | GU: 7.5 (4.3)<br>UU: 9.92 (7.74) | GU: 22.4 (6.35)<br>UU: 2.7 (1.91), MMSE |

a, modified version; b, percentage of persons with a MMSE score <24; c, MMSE < 19 points; CNA, nine-item care needs assessment; d, median; GU, geriatric unit; MMF, mild-to-moderate frailty; MMS, Mini-Mental State Examination; NR, not reported; PF, pre-frail; RB, robust; SF, severe frailty; SPMSQ, Short Portable Mental Status Questionnaire; UU, usual unit.

### Outcome results

The calculated and estimated incidences of HAD are shown in Table 2, categorized per ADL task and set of tasks. The pooled incidence of HAD for the overall score included two studies reporting the Katz Index of ADLs 1963 [23, 27] and two others used the Katz Index of ADLs 1970 [17, 21] for a total of 4,020 patients. Figure 1 presents the pooled incidence of HAD (total score) of 37% (95% CI 0.31–0.42). Heterogeneity was substantial at 90%. The risk of bias of the four studies raised some concerns.

**Table 2.**
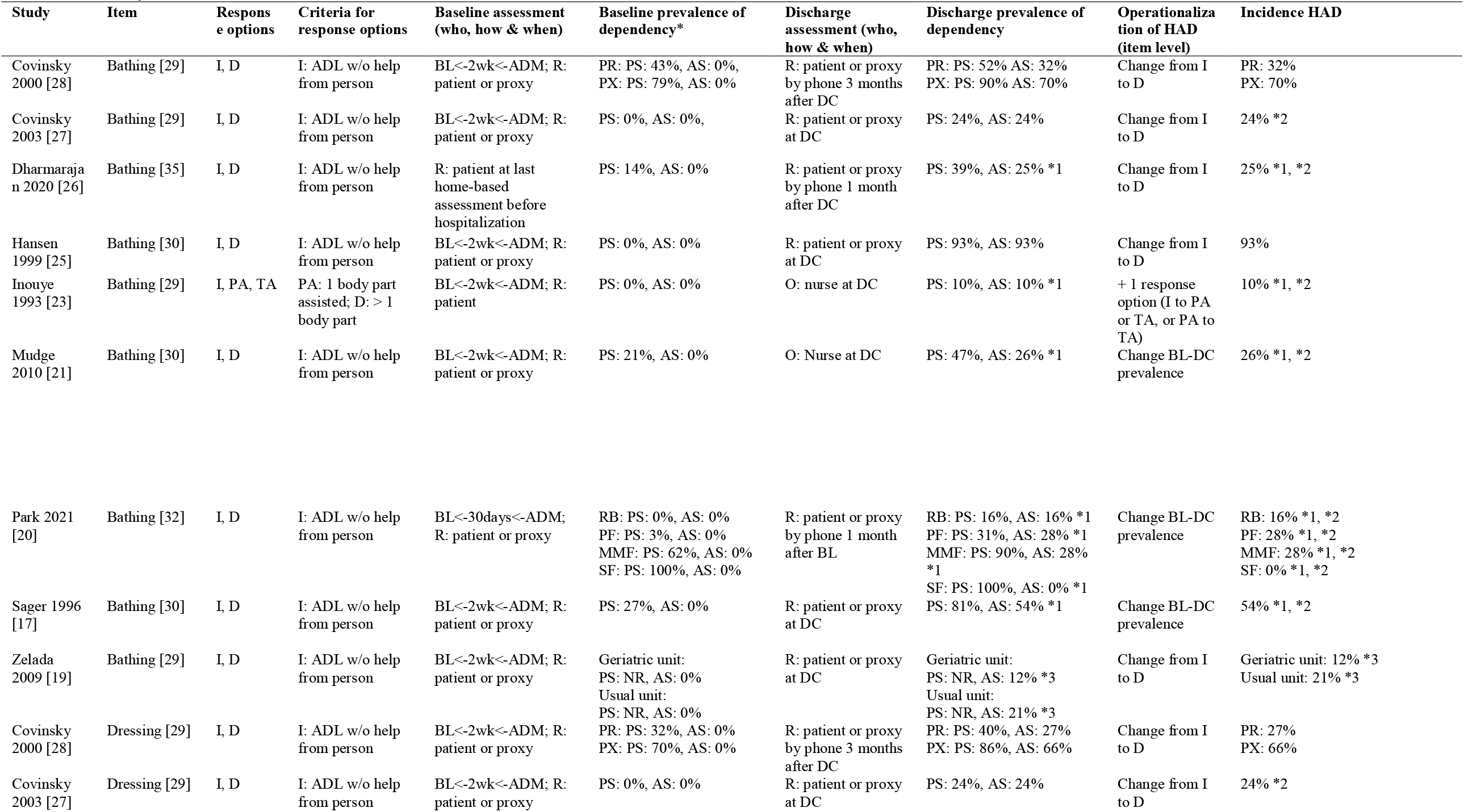
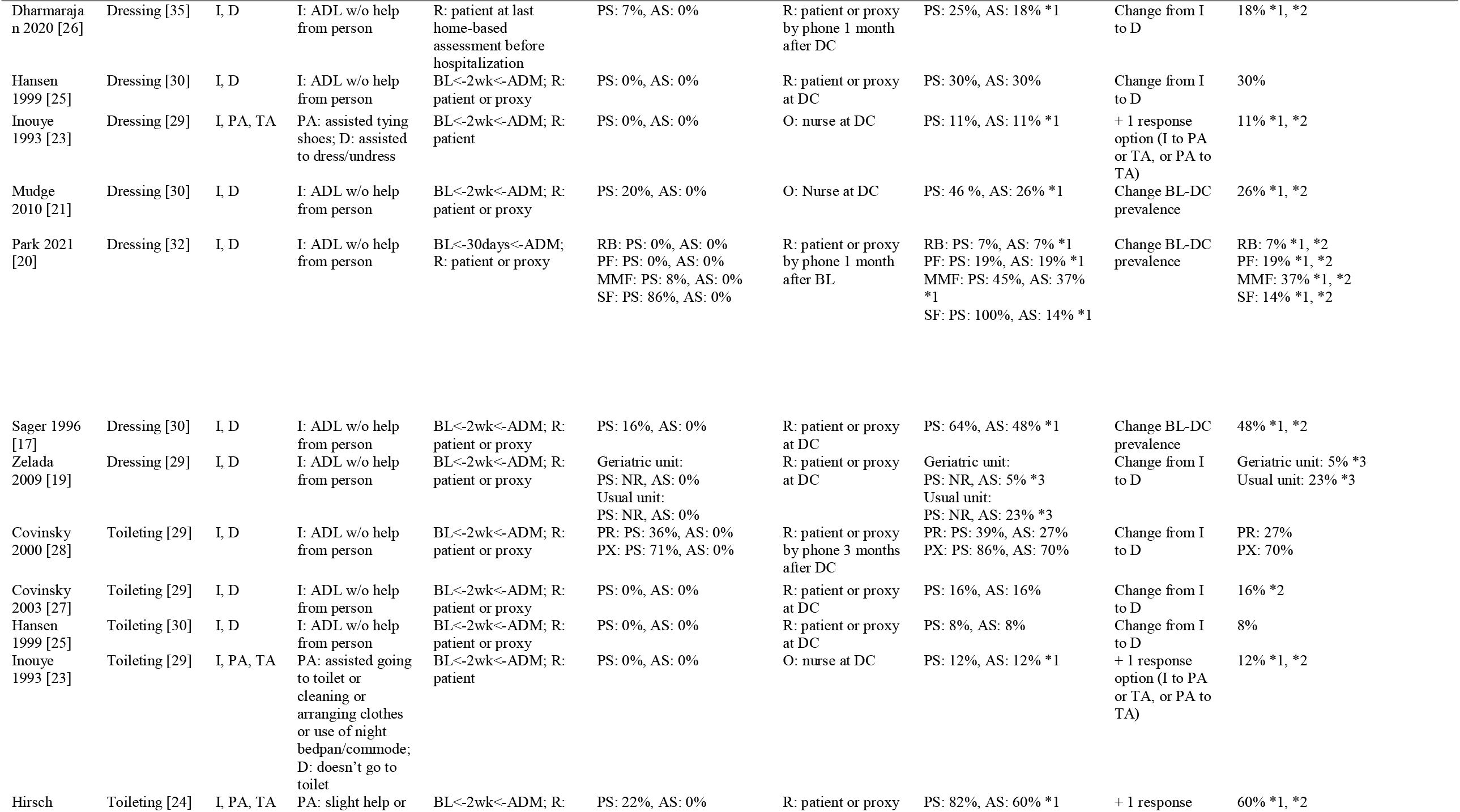
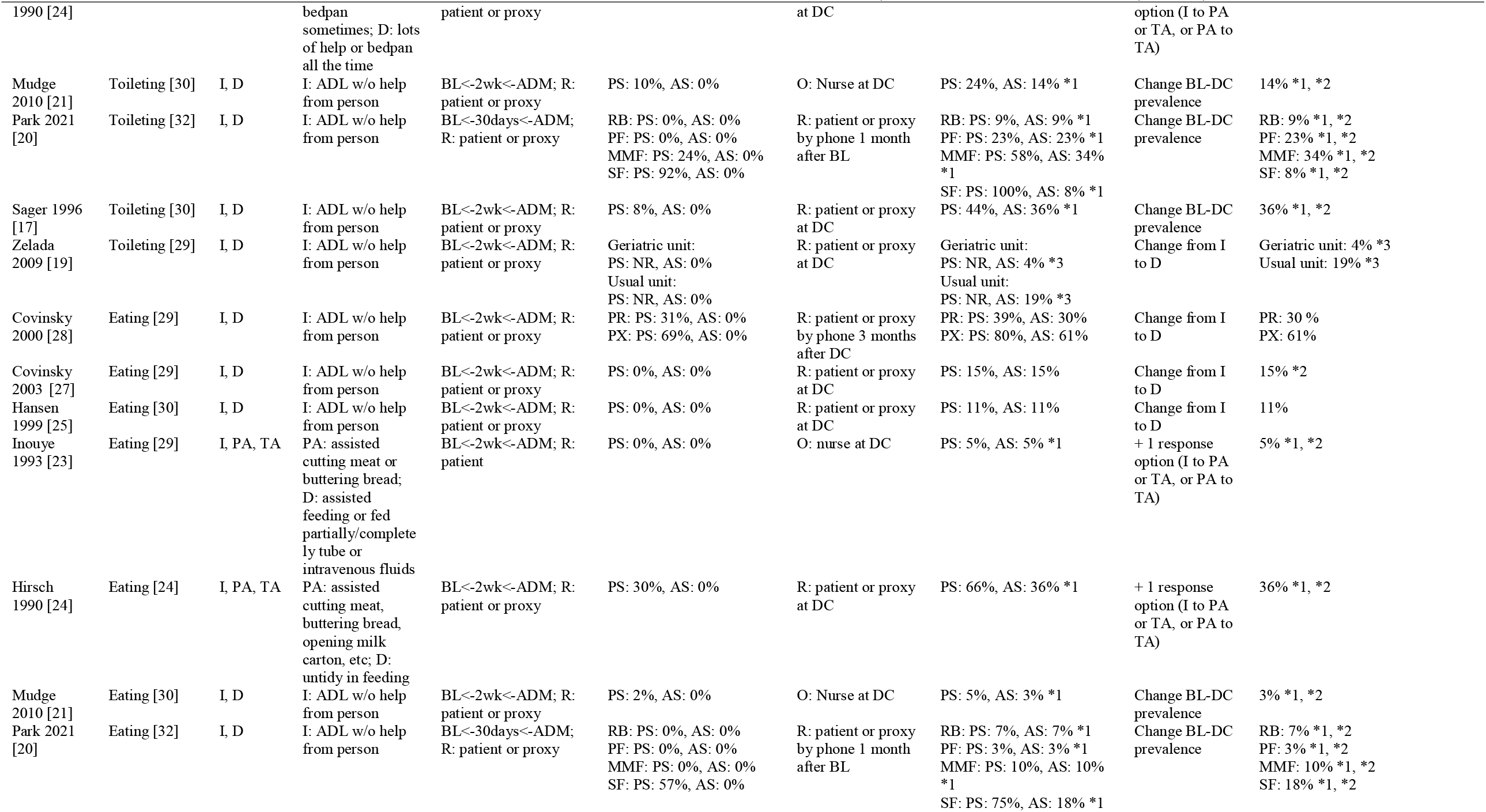
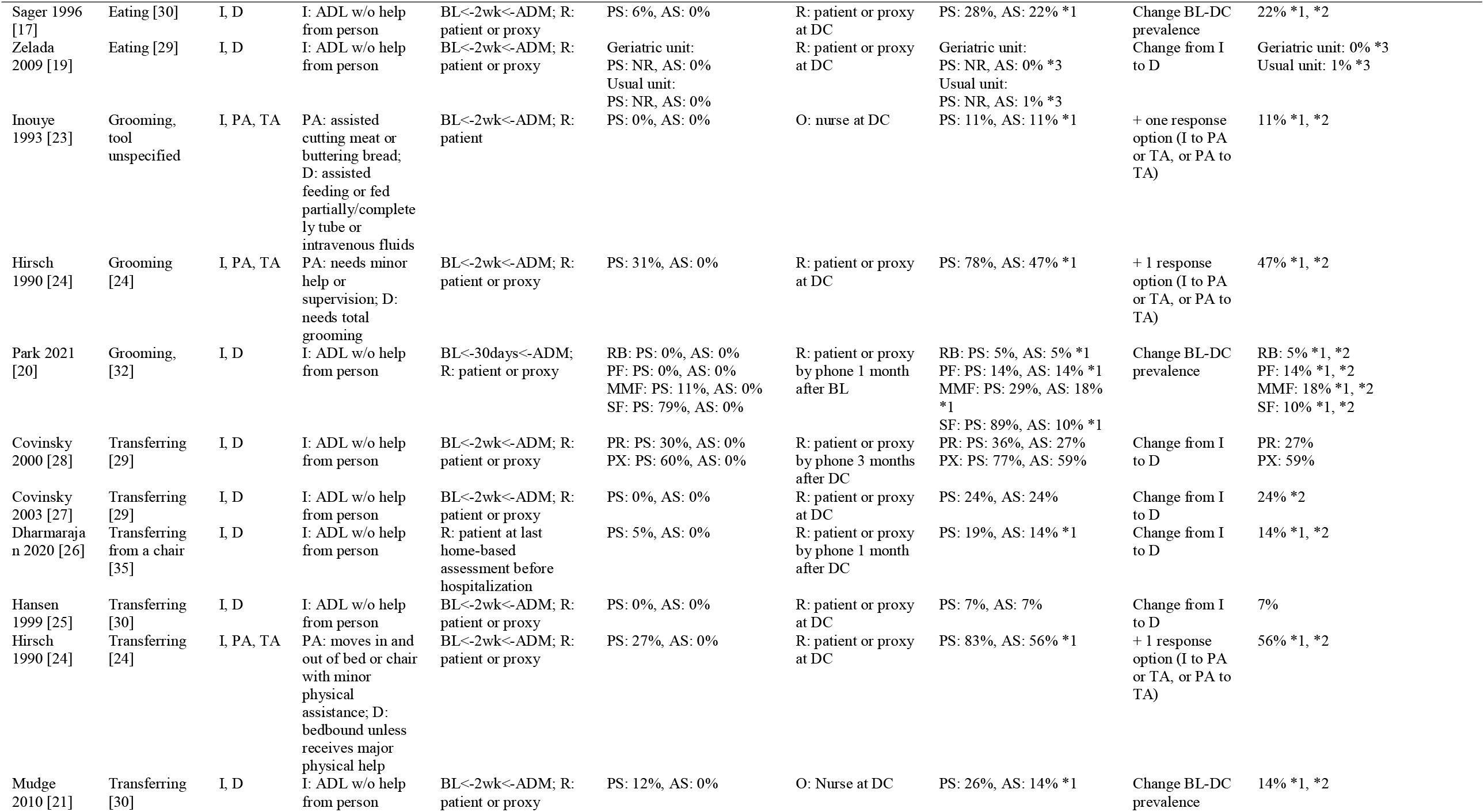
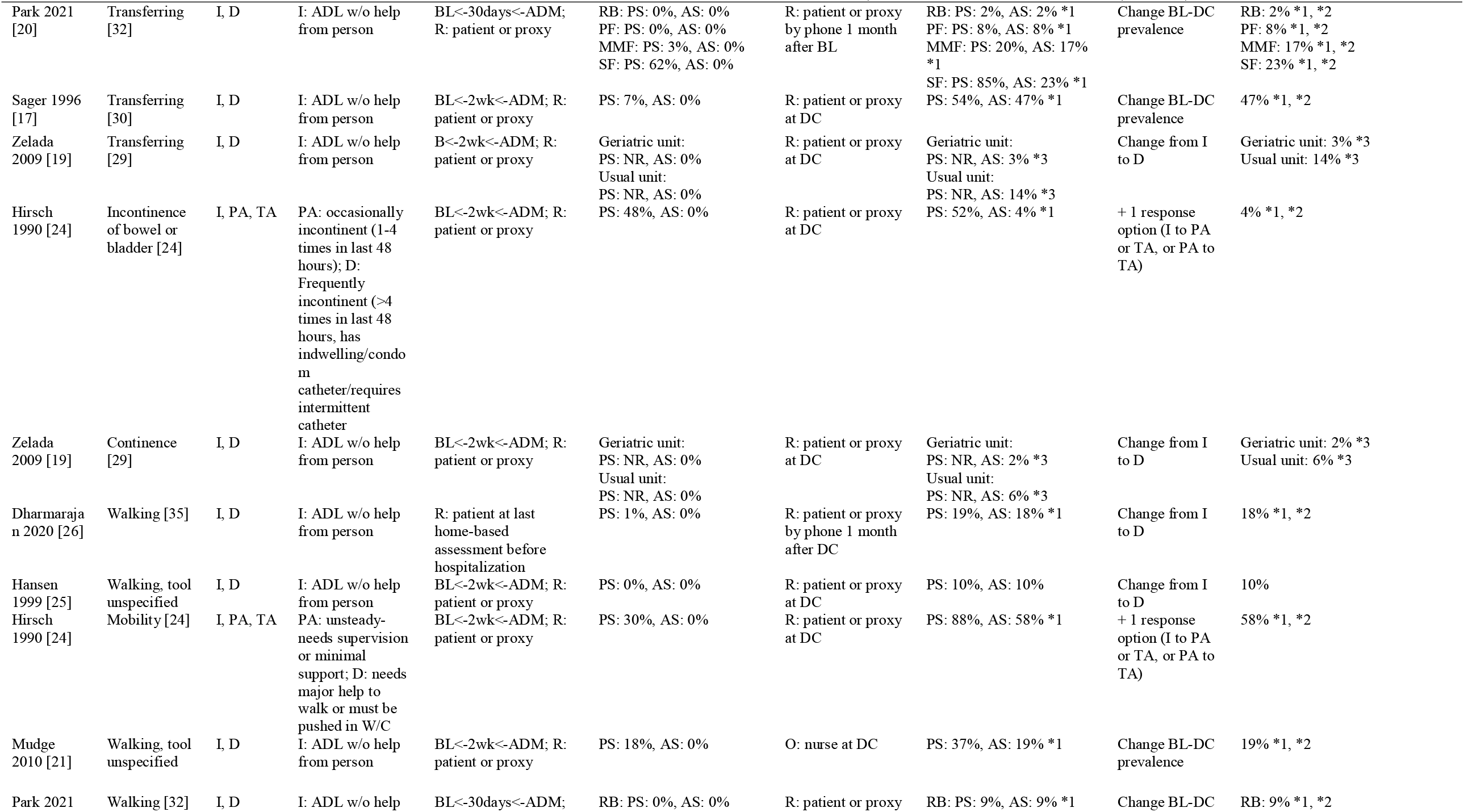
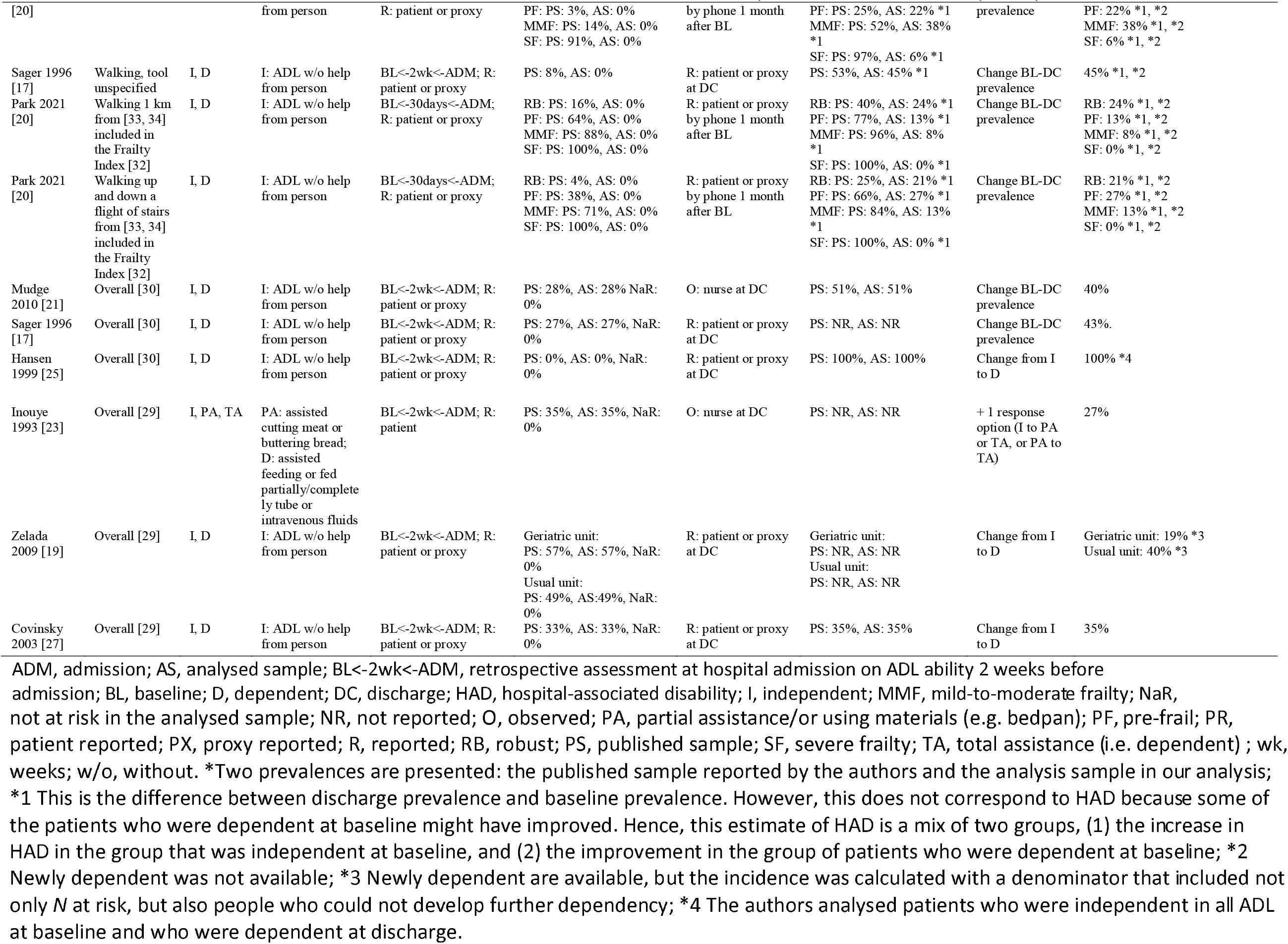
Comparison of incidence items

**Figure 1.**
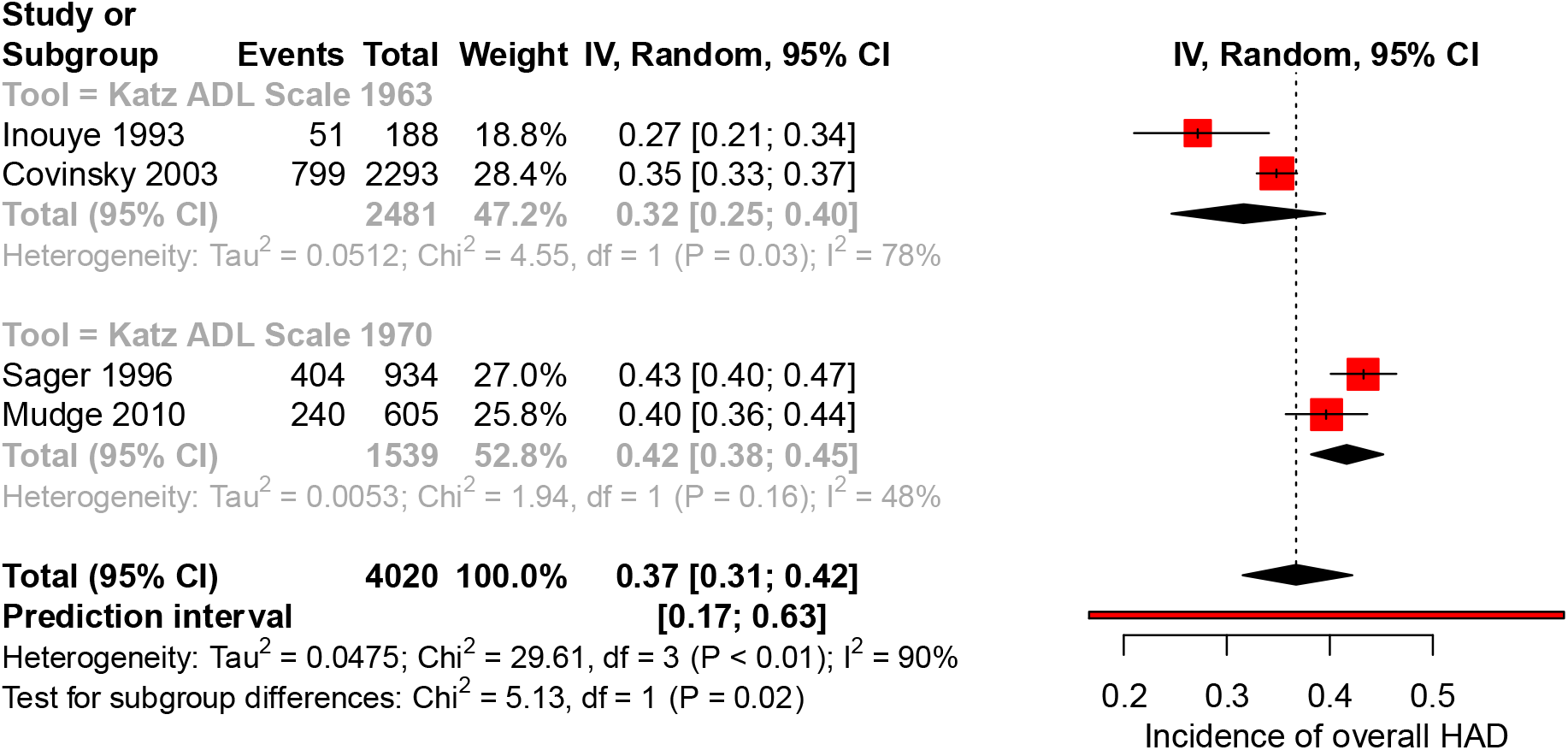
Forest plot of the pooled results of overall incidence of hospital-associated disability (HAD).

Table 2 shows the results of the incidences at the item level. Adequate data for the incidence calculation at the item level was reported only by two studies [25, 28]. The incidence of HAD for the individual items was in the range 32–93% for bathing, 27–66% for dressing, 8–70% for toileting, 11–61% for eating, 7–59% for transferring, and 10% for walking. For the other studies, it was not possible to calculate the incidence of HAD (either total or single-item level) due to insufficient data. One RCT could not be integrated into Table 2 because the authors reported only the mean change in ADL score, and the results of this study were described narratively [22]. The control group worsened in all items of ADL, negative values indicating a decline in the Barthel mean change score. The three items with the highest mean change score from baseline to discharge were: transferring (–2.06 points; 95% CI –1.44 to –2.71), climbing stairs (–1.36 points; 95% CI –0.84 to –1.91), and toileting (– 1.23 points; 95% CI –0.76 to –1.69). The score for these individual tasks ranged from 0 points (dependent) to independent with a maximum of 15 points for transferring, 10 points for climbing stairs and 10 points for using the toilet.

The risk of bias of all studies presented some concerns.

Eleven studies were included in the identification of the tools or functional tasks used to assess ADL in hospitalized older patients and in the ranking of the most sensitive task to detect changes in disability [17, 19-28]. The review identified five assessment tools, two sets of tasks, and individual items assessing ADL: Katz Index of ADLs 1963 [29], Katz Index of ADLs 1970 [30], Barthel Index [31], 8 items from the nine-item Care Needs Assessment tool [24], 7 ADL items form the Frailty Index [32] and 2 activities from the Nagi and Rosow-Breslau scales [33, 34] integrated in the Frailty Index [32], and individual items proposed by Gill [35] (bathing, dressing, transferring and walking across the room). Walking was assessed in two studies [21, 25], while one other study [23] assessed the task of grooming by referring to the Katz Index of ADLs. However, the item walking and grooming were not in the original version of the Katz Index, and were considered separately.

Table 3 shows the number of studies that reported the tasks (items) and the ranking of the most sensitive tasks for detecting changes in disability.

**Table 3.** Ranking of the most sensitive tasks or sets of tasks for detecting change in disability

| Number of sub-groups reporting this item | Item | Mean rank |
| --- | --- | --- |
| 7 | Overall | 1.57 |
| 1 | Mobility | 2 |
| 14 | Bathing | 2.5 |
| 14 | Dressing | 3.07 |
| 14 | Toileting | 3.86 |
| 1 | Transferring from a chair | 4 |
| 8 | Walking | 4 |
| 4 | Walking up and down a flight of stairs | 5 |
| 13 | Transferring | 5.15 |
| 6 | Grooming | 5.17 |
| 14 | Eating | 5.79 |
| 2 | Continence | 6 |
| 1 | Incontinence of bowel or bladder | 6 |
| 2 | Walking 1 km | 6.25 |

### Methodological quality

Figure 2 presents a summary plot of the assessment of methodological quality using the JBI checklist. All of the included studies and study sub-groups reported an adequate sample frame and recruitment procedure, and appropriately detailed the study subjects and setting. Approximately 37% of the included studies reported an adequate study sample size, and for approximately 63% it was inadequate. All of the sub-groups described the study subjects and setting well. Adequate analysis with sufficient coverage of the identified sample was conducted in approximately 25% of studies and was unclear in 75% of studies. All study sub-groups were unclear regarding the methods used for identification of the condition. The participant’s condition was measured in a standard and reliable way in approximately 37% of the sub-groups and was unclear in 63%. The statistical analysis was adequate in approximately 19% and inadequate in 81% of the included studies. The response rate of the subgroups was good in 13%, unclear in 6%, and inadequate in 81% of the studies under evaluation. The rating of the individual items for each study is available in the supplementary material (Appendix 3).

**Figure 2.**
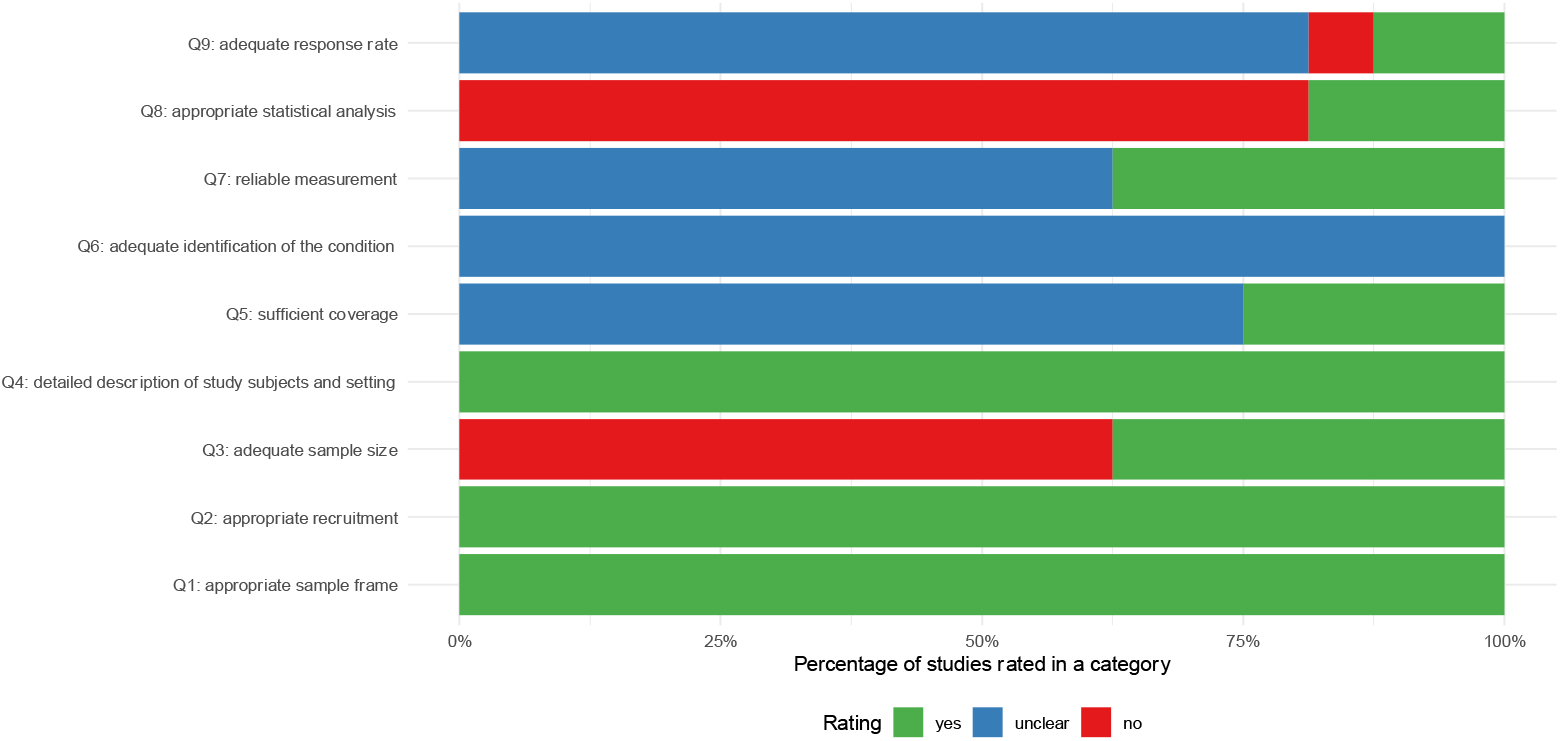
Joanna Briggs Institute (JBI) checklist summary plot.

## Discussion

The aims of this systematic review were to evaluate the incidence of HAD, to identify the tools or sets of tasks used in a hospital setting to assess ADL in older people, and to determine which functional task is most sensitive for detecting changes in disability.

The pooled incidence of HAD (total score) was 37%. The difference in incidence according to the two versions of the Katz Index of ADLs was 8%. This difference could be explained by the fact that patients in the studies by Sager [17] and Mudge [21] were more dependent at baseline compared with the volunteers included in the studies by Inouye [23] and Covinsky [27]. The difference could not be explained by comorbidities (e.g. with the APACHE II score or the Charlson Comorbidity Index) as the authors of the studies by Sager and Mudge did not report these values.

Loyd et al. [9] found a prevalence of HAD of 30% in older adults hospitalized for acute care. The current review focused on the number of new cases of HAD, i.e. the incidence of HAD. This latter measure considers the number of new cases of decline over the population at risk of developing a new decline. To our knowledge, no other review has investigated the overall incidence of HAD and the incidence at the item level in a population of older patients over 65 years hospitalized for acute care. In general, the incidence of HAD at the item level was higher when disability was reported by the surrogates than when it was reported by patients. Similar results were found in previous research [36].

Regarding the tools or sets of tasks used in a hospital setting to assess ADL in older people, this review identified that the Katz Index of ADLs was the most reported tool in the included studies. The Barthel Index and the Katz Index of ADLs are the oldest tools used for assessing ADL [37]. Seven studies used the Katz Index of ADLs, and none of them assessed the continence item that was include in the original version [17, 21, 23, 25, 27, 28]. In addition, the Katz Index of ADLs was modified by adding another item of walking in three studies [17, 21, 25] and adding the item grooming in one studies [23].

This review found that the most sensitive task or set of tasks for detecting changes in disability was use of the total score, which is a composite score of several tasks. However, this composite score was based on a different set of tasks in individual studies. For example, continence was only included in the total score in Zelada et al.’s study [19]. The four next most sensitive tasks for detecting changes in disability were mobility, bathing, dressing, and toileting. It should be noted that items related to mobility and transfer from a chair appeared relatively high in the rankings and were assessed based on only one study. It was observed that the tasks that are generally considered difficult were not ranked as the most sensitive to change. For example, the item walking 1 km may be considered by many older patients as difficult, but this item did not seem very sensitive to change. This observation might be explained by the fact that many subjects were already dependent on help to perform this task and did not have the potential to decline further, hence resulting in a low incidence. The walking item was generally poorly described and was therefore categorized separately from the mobility item due to a lack of information about the similarity of these two items and their judging criteria. The item walking 1 km was considered more difficult than the walking task, which usually refers to a smaller distance; hence these functional tasks were categorized separately. This has the disadvantage of resulting in fewer studies per functional task.

To determine which was the most sensitive task or set of tasks for detecting changes in disability, a meta-analysis of incidences would have been the preferred method. However, due to heterogeneity between studies and missing data it was not possible to pool the data. Therefore, the mean of the ranking was analysed, and this analysis should be interpreted with caution. Futures studies may change the ranking.

An overestimation of HAD might be present for two main reasons. First, differences in the execution of the ADL assessment might have resulted in overestimation of the HAD incidence. In two studies [21, 23], the baseline value for ADL ability was asked retrospectively to patients at admission, while ADL ability at discharge was assessed by an experienced healthcare professional. Older patients tend to overestimate their ADL ability [38, 39], their ability to step over an obstacle [40] and their motor performance [41]. Kawasaki and Tozawa hypothesized that the patients’ overestimation of their physical or functional capacities might be explained by the absence of recognition of their decline in motor performance [41]. Observer-based assessments of ADL tasks by healthcare professionals were more accurate than patients’ self-reported ADL values [42, 43].

Furthermore, this review included studies investigating pathologies that should not have a long-lasting effect on functional disability. However, it seems that there might be a combination of HAD and disease-related disability. Covinsky et al.’s study [27] was the only one to present the disability trajectories of each patient group and their development over time. These authors reported the percentage of patients whose ADL values declined between baseline (two weeks before hospitalization) and hospital admission (which is related to the condition) and those who had not recovered by discharge.

We believe that this combination of HAD and disease-related disability may also have contributed to an overestimation of HAD. In general, the studies did not separately report the numbers of persons with a disease-related disability and those with HAD, but only reported a combined total. Hence, it is unclear whether the failure to recover is due to hospitalization.

Moreover, the current ADL assessment tools present measurement properties limitations. For example, the Barthel Index showed a floor [44] and ceiling effect [44-46]. Two of the studies included in the current review reported a ceiling effect of the Katz Index of ADLs [17, 21].

A strength of the current study is that, to the best of our knowledge, this is the first review to investigate the incidence of HAD in a population of older patients over 65 years of age, hospitalized for acute care. To our knowledge, previous systematic reviews did not assess the incidence of HAD at the item level.

The study has a number of limitations. The screening process was performed independently by two reviewers for only 20% of the records. However, we believe that this has only limited negative influence, as the agreement between the reviewers was high (above the predefined kappa coefficient of agreement of 0.8) and this procedure is accepted and suggested in rapid systematic review [10]. Another limitation is that we could not conduct a meta-analysis for the individual tasks due to insufficient data. With our reported methods, the certainty in the presented estimated incidence of HAD at the item level is very low. The estimated incidence, calculated as the difference between the discharge and baseline prevalence, should be interpreted with caution, as this does not consider the change over time of independent and dependent patients from baseline to discharge. The prevalence of disability at discharge does not distinguish between: (i) those who remain dependent between baseline and discharge, (ii) the newly dependent, and (iii) those who became independent at discharge. Therefore, these estimated incidences cannot be considered true incidence. In addition, small sample size bias cannot be totally excluded in the current review.

### Implications and further research

Further studies should investigate the reasons for the overestimation of HAD. As reported above, there is a need to develop a more sensitive tool that reflects the true functional status of older patients (over 65 years) before hospitalization for acute care. The systematic integration of proxies in the evaluation of functional status before hospitalization, in addition to the patient self-reported assessment, needs to be deepened. Regarding the implication for healthcare professionals, activities such as mobility, bathing, and dressing seem to be the most sensitive to detect changes in disability. The development of targeted training interventions for these ADL activities should be considered in future studies. HAD is a relevant problem, and a systematic appraisal of existing intervention studies addressing this problem is missing. Future research should consider interview methods to help patients better remember their abilities at home in order to reflect their true ability in ADL function.

## Conclusion

Functional decline in older patients over 65 years of age, due to hospitalization for acute care, is an important problem, with an incidence of 37% based on the overall score of ADL assessment. This incidence might be overestimated, due to a combination of disease-related disability and HAD, while measurement tools may also present some limitations. Furthermore, it is not possible to draw a definitive conclusion on the incidence of HAD at the item level, as there is insufficient data reported to enable the results of individual tasks to be pooled. This systematic review found that the most relevant ADL tasks for detecting disability are mobility, bathing, and dressing, although this result should be interpreted with caution. Further studies should investigate the overestimation of HAD and how to overcome this limitation.

## Data Availability

All data produced in the present work are contained in the manuscript

## Registration and protocol

The protocol of this rapid systematic review has not been published in a peer-review journal. However, it is registered at OSF registries (https://osf.io/9jez4/) identifier: DOI 10.17605/OSF.IO/9JEZ4.

## APPENDICES

### Appendix 1. Search strategy

**Appendix 1.1.** Medline (via Ovid) search.

| Number | Query | Results<br>the 26<br>Aug 2021 |
| --- | --- | --- |
| 1 | Hospitalisation.ti,ab. | 17,353 |
| 2 | exp *Hospitalization/ | 93,562 |
| 3 | hospitalization.ti,ab. | 137,158 |
| 4 | Hospital.ti,ab. | 1,075,670 |
| 5 | Hospitalized.ti,ab. | 112,702 |
| 6 | Hospitalised.ti,ab. | 13,156 |
| 7 | In-Hospital.ti,ab. | 98,939 |
| 8 | Inhospital.ti,ab. | 1,870 |
| 9 | Inpatient.ti,ab. | 88,351 |
| 10 | 1 or 2 or 3 or 4 or 5 or 6 or 7 or 8 or 9 | 1,300,482 |
| 11 | functional.ti,ab. | 1,338,546 |
| 12 | Disability.ti,ab. | 168,863 |
| 13 | disabled.ti,ab. | 24,221 |
| 14 | Activity of daily living.ti,ab. | 2,185 |
| 15 | Activities of daily living.ti,ab. | 28,860 |
| 16 | Activity of daily life.ti,ab. | 482 |
| 17 | Activities of daily life.ti,ab. | 1,403 |
| 18 | exp *"Activities of Daily Living"/ | 45,453 |
| 19 | activities.ti,ab. | 755,046 |
| 20 | daily.ti,ab. | 557,713 |
| 21 | living.ti,ab. | 373,448 |
| 22 | 19 and 20 and 21 | 32,687 |
| 23 | Activity.ti,ab. | 2,802,780 |
| 24 | daily.ti,ab. | 557,713 |
| 25 | living.ti,ab. | 373,448 |
| 26 | 23 and 24 and 25 | 8,758 |
| 27 | Participation.ti,ab. | 159,573 |
| 28 | Activity.ti,ab. | 2,802,780 |
| 29 | limitation.ti,ab. | 90,604 |
| 30 | 28 and 29 | 11,618 |
| 31 | 11 or 12 or 13 or 14 or 15 or 16 or 17 or 18 or 22 or 26 or 27 or 30 | 1,699,597 |
| 32 | prevalence.ti,ab. | 683,571 |
| 33 | frequency.ti,ab. | 878,298 |
| 34 | burden.ti,ab. | 230,392 |
| 35 | exp Random Allocation/ | 105,786 |
| 36 | random*.ti,ab. | 1,247,309 |
| 37 | frequencies.ti,ab. | 197,525 |
| 38 | inciden*.ti,ab. | 951,438 |
| 39 | incidence.ti,ab. | 805,068 |
| 40 | risk.ti,ab. | 2,320,682 |
| 41 | hazard ratio.ti,ab. | 108,340 |
| 42 | hazard ratios.ti,ab. | 29,363 |
| 43 | odds.ti,ab. | 363,083 |
| 44 | chance.ti,ab. | 69,946 |
| 45 | percent.ti,ab. | 295,410 |
| 46 | percentage.ti,ab. | 403,689 |
| 47 | percentages.ti,ab. | 66,053 |
| 48 | rate.ti,ab. | 2,216,830 |
| 49 | rates.ti,ab. | 1,158,092 |
| 50 | occurrence.ti,ab. | 386,376 |
| 51 | responsiveness.ti,ab. | 105,969 |
| 52 | sensitivity to change.ti,ab. | 2,604 |
| 53 | 32 or 33 or 34 or 35 or 36 or 37 or 38 or 39 or 40 or 41 or 42 or 43 or 44 or 45 or 46 or 47 or 48 or 49 or 50 or 51 or 52 | 7,901,067 |
| 54 | older.ti,ab. | 474,685 |
| 55 | elderly.ti,ab. | 259,926 |
| 56 | geriatric.ti,ab. | 44,737 |
| 57 | exp *Aged/ | 26,580 |
| 58 | aged.ti,ab. | 623,417 |
| 59 | frail.ti,ab. | 13,694 |
| 60 | frails.ti,ab. | 13 |
| 61 | exp *Frailty/ | 4,180 |
| 62 | frailty.ti,ab. | 16,886 |
| 63 | frailness.ti,ab. | 20 |
| 64 | ageing.ti,ab. | 45,000 |
| 65 | senior.ti,ab. | 36,394 |
| 66 | elders.ti,ab. | 9,003 |
| 67 | 54 or 55 or 56 or 57 or 58 or 59 or 60 or 61 or 62 or 63 or 64 or 65 or 66 | 1,256,648 |
| 68 | hospital-associated.ti,ab. | 1,276 |
| 69 | hospital associated.ti,ab. | 1,276 |
| 70 | hospital-acquired.ti,ab. | 10,652 |
| 71 | hospital acquired.ti,ab. | 10,652 |
| 72 | hospital-related.ti,ab. | 806 |
| 73 | hospital related.ti,ab. | 806 |
| 74 | hospitalization-associated.ti,ab. | 309 |
| 75 | hospitalization associated.ti,ab. | 309 |
| 76 | hospitalisation-associated.ti,ab. | 38 |
| 77 | hospitalisation associated.ti,ab. | 38 |
| 78 | hospitalization-acquired.ti,ab. | 3 |
| 79 | hospitalization acquired.ti,ab. | 3 |
| 80 | hospitalisation-acquired.ti,ab. | 1 |
| 81 | hospitalisation acquired.ti,ab. | 1 |
| 82 | hospitalization-related.ti,ab. | 220 |
| 83 | hospitalization related.ti,ab. | 220 |
| 84 | hospitalisation-related.ti,ab. | 28 |
| 85 | hospitalisation related.ti,ab. | 28 |
| 86 | before hospitalisation.ti,ab. | 105 |
| 87 | before hospitalization.ti,ab. | 1,090 |
| 88 | after hospitalisation.ti,ab. | 434 |
| 89 | after hospitalization.ti,ab. | 3,890 |
| 90 | (functional deterioration adj3 hospital*).ti,ab. | 20 |
| 91 | (hospital* adj3 functional loss).ti,ab. | 13 |
| 92 | (functional status adj3 Hospital*).ti,ab. | 342 |
| 93 | (los* adj3 independence).ti,ab. | 1,369 |
| 94 | (exposure adj2 hospital*).ti,ab. | 797 |
| 95 | (functional decline adj4 hospital*).ti,ab. | 313 |
| 96 | 68 or 69 or 70 or 71 or 72 or 73 or 74 or 75 or 76 or 77 or 78 or 79 or 80 or 81 or 82 or 83 or 84 or 85 or 86 or 87 or 88 or 89 or 90 or 91 or 92 or 93 or 94 or 95 | 21,232 |
| 97 | 10 and 31 and 53 and 67 and 96 | 740 |

**Appendix 1.2.** Embase search.

| Number | Query | Results the<br>26 Aug 2021 |
| --- | --- | --- |
| 1 | hospitalisation:ti,ab | 28,806 |
| 2 | 'hospitalization'/exp | 421,223 |
| 3 | hospitalization:ti,ab | 230,090 |
| 4 | hospital:ti,ab | 1,708,899 |
| 5 | hospitalized:ti,ab | 175,158 |
| 6 | hospitalised:ti,ab | 20,314 |
| 7 | 'in hospital':ti,ab | 159,194 |
| 8 | inhospital:ti,ab | 6,247 |
| 9 | inpatient:ti,ab | 151,612 |
| 10 | #1 OR #2 OR #3 OR #4 OR #5 OR #6 OR #7 OR #8 OR #9 | 2,125,565 |
| 11 | functional:ti,ab | 1,651,619 |
| 12 | disability:ti,ab OR disabled:ti,ab | 268,385 |
| 13 | 'activity of daily living':ti,ab | 3,263 |
| 14 | 'activities of daily living':ti,ab | 40,031 |
| 15 | 'activity of daily life':ti,ab | 341 |
| 16 | 'activities of daily life':ti,ab | 1,857 |
| 17 | 'daily life activity'/exp/mj | 14,822 |
| 18 | participation:ti,ab | 207,496 |
| 19 | activity:ti,ab | 3,491,683 |
| 20 | limitation:ti,ab | 120,101 |
| 21 | #19 AND #20 | 16,085 |
| 22 | #11 OR #12 OR #13 OR #14 OR #15 OR #16 OR #17 OR #18 OR #21 | 2,131,790 |
| 23 | prevalence:ti,ab | 964,018 |
| 24 | frequency:ti,ab | 1,138,741 |
| 25 | burden:ti,ab | 352,672 |
| 26 | 'randomized controlled trial'/exp/mj | 11,937 |
| 27 | 'randomization'/exp | 91,820 |
| 28 | inciden*:ti,ab | 1,362,309 |
| 29 | incidence:ti,ab | 1,157,883 |
| 30 | risk:ti,ab | 3,334,654 |
| 31 | 'hazard ratio':ti,ab | 154,212 |
| 32 | 'hazard ratios':ti,ab | 45,812 |
| 33 | odds:ti,ab | 460,665 |
| 34 | chance:ti,ab | 101,595 |
| 35 | percent:ti,ab | 402,184 |
| 36 | percentage:ti,ab | 577,947 |
| 37 | percentages:ti,ab | 92,283 |
| 38 | rate:ti,ab | 3,018,423 |
| 39 | rates:ti,ab | 1,571,532 |
| 40 | occurrence:ti,ab | 499,939 |
| 41 | responsiveness:ti,ab | 131,335 |
| 42 | 'sensitivity to change':ti,ab | 3,234 |
| 43 | frequencies:ti,ab | 237,267 |
| 44 | #23 OR #24 OR #25 OR #26 OR #27 OR #28 OR #29 OR #30 OR #31 OR #32 OR #33 OR #34 OR #35 OR #36 OR #37 OR #38 OR #39 OR #40OR #41 OR #42 OR #43 | 10,403,809 |
| 45 | older:ti,ab | 653,439 |
| 46 | elderly:ti,ab | 367,177 |
| 47 | geriatric:ti,ab | 69,535 |
| 48 | 'aged'/exp | 3,321,306 |
| 49 | aged:ti,ab | 854,083 |
| 50 | frail:ti,ab | 21,314 |
| 51 | frails:ti,ab | 28 |
| 52 | frailty:ti,ab | 25,936 |
| 53 | 'frailty'/exp | 15,719 |
| 54 | frailness:ti,ab | 29 |
| 55 | ageing:ti,ab | 60,209 |
| 56 | senior:ti,ab | 51,231 |
| 57 | elders:ti,ab | 11,925 |
| 58 | #45 OR #46 OR #47 OR #48 OR #49 OR #50 OR #51 OR #52 OR #53 OR #54 OR #55 OR #56 OR #57 | 5,394,076 |
| 59 | 'hospital associated':ti,ab | 1,830 |
| 60 | 'hospital-associated':ti,ab | 1,836 |
| 61 | 'hospital acquired':ti,ab | 15,633 |
| 62 | 'hospital-acquired':ti,ab | 15,725 |
| 63 | 'hospital related':ti,ab | 1,235 |
| 64 | 'hospital-related':ti,ab | 1,241 |
| 65 | 'hospitalization associated':ti,ab | 442 |
| 66 | 'hospitalization-associated':ti,ab | 444 |
| 67 | 'hospitalisation-associated':ti,ab | 58 |
| 68 | 'hospitalisation associated':ti,ab | 58 |
| 69 | 'hospitalization acquired':ti,ab | 4 |
| 70 | 'hospitalization-acquired':ti,ab | 4 |
| 71 | 'hospitalisation acquired':ti,ab | 0 |
| 72 | 'hospitalisation-acquired':ti,ab | 0 |
| 73 | 'hospitalization related':ti,ab | 394 |
| 74 | 'hospitalization-related':ti,ab | 396 |
| 75 | 'hospitalisation related':ti,ab | 45 |
| 76 | 'hospitalisation-related':ti,ab | 45 |
| 77 | 'before hospitalisation':ti,ab | 202 |
| 78 | 'before hospitalization':ti,ab | 1,692 |
| 79 | 'after hospitalisation':ti,ab | 671 |
| 80 | 'after hospitalization':ti,ab | 6,053 |
| 81 | functional:ti,ab AND<br>((deterioration NEAR/3 hospital*):ti,ab) | 83 |
| 82 | ((hospital* NEAR/3 functional):ti,ab) AND loss:ti,ab | 241 |
| 83 | functional:ti,ab AND ((status NEAR/3 hospital*):ti,ab) | 859 |
| 84 | (los* NEAR/3 independence):ti,ab | 2,106 |
| 85 | (exposure NEAR/2 hospital*):ti,ab | 1,197 |
| 86 | functional:ti,ab AND ((decline NEAR/4 hospital*):ti,ab) | 582 |
| 87 | #59 OR #60 OR #61 OR #62 OR #63 OR #64 OR #65 OR<br>#66 OR #67 OR #68 OR #69 OR #70 OR #71 OR #72 OR<br>#73 OR #74 OR #75 OR #76OR #77 OR #78 OR #79 OR<br>#80 OR #81 OR #82 OR #83 OR #84 OR #85 OR #86 | 21,314 |
| 88 | #10 AND #22 AND #44 AND #58 AND #87 | 1,557 |

**Appendix 1.3.** Cochrane search.

| ID | Search | Results the<br>26 Aug 2021 |
| --- | --- | --- |
| 1 | hospitalisation:ti,ab | 30,002 |
| 2 | MeSH descriptor: [Hospitalization] explode all trees | 14,616 |
| 3 | hospitalization:ti,ab | 29,933 |
| 4 | hospital:ti,ab | 129,129 |
| 5 | hospitalized:ti,ab | 16,701 |
| 6 | hospitalised:ti,ab | 16,701 |
| 7 | in-hospital:ti,ab | 12,291 |
| 8 | inhospital:ti,ab | 5,816 |
| 9 | inpatient:ti,ab | 12,747 |
| 10 | #1 OR #2 OR #3 OR #4 OR #5 OR #6 OR #7 OR #8 OR #9 | 167,548 |
| 11 | functional:ti,ab | 76,938 |
| 12 | disability:ti,ab | 31,499 |
| 13 | disabled:ti,ab | 1,712 |
| 14 | "activity of daily living":ti,ab | 711 |
| 15 | "activities of daily living":ti,ab | 7,311 |
| 16 | "activity of daily life":ti,ab | 88 |
| 17 | "activities of daily life":ti,ab | 338 |
| 18 | MeSH descriptor: [Activities of Daily Living] explode all trees | 9,739 |
| 19 | participation:ti,ab | 30,692 |
| 20 | activity:ti,ab | 132,423 |
| 21 | limitation:ti,ab | 7,562 |
| 22 | #20 AND #21 | 1,315 |
| 23 | #11 OR #12 OR #13 OR #14 OR #15 OR #16 OR #17 OR #18 OR #19 OR #22 | 139,849 |
| 24 | prevalence:ti,ab | 33,961 |
| 25 | frequency:ti,ab | 79,109 |
| 26 | burden:ti,ab | 22,207 |
| 27 | MeSH descriptor: [Random Allocation] explode all trees | 20,643 |
| 28 | MeSH descriptor: [Randomized Controlled Trial] explode all trees | 119 |
| 29 | inciden*:ti,ab | 125,517 |
| 30 | incidence:ti,ab | 116,076 |
| 31 | risk:ti,ab | 219,370 |
| 32 | "hazard ratio":ti,ab | 22,039 |
| 33 | "hazard ratios":ti,ab | 3,476 |
| 34 | odds:ti,ab | 27,403 |
| 35 | chance:ti,ab | 8,943 |
| 36 | percent:ti,ab | 34,597 |
| 37 | percentage:ti,ab | 50,616 |
| 38 | percentages:ti,ab | 6,400 |
| 39 | rate:ti,ab | 282,297 |
| 40 | rates:ti,ab | 136,486 |
| 41 | occurrence:ti,ab | 27,342 |
| 42 | responsiveness:ti,ab | 7,597 |
| 43 | "sensitivity to change":ti,ab | 357 |
| 44 | frequencies:ti,ab | 7,361 |
| 45 | #24 OR #25 OR #26 OR #27 OR #28 OR #29 OR #30 OR #31 OR #32 OR #33 OR #34 OR #35 OR #36 OR #37 OR #38 OR #39 OR #40 OR #41 OR #42 OR #43 OR #44 | 728,891 |
| 46 | older:ti,ab | 59,544 |
| 47 | elderly:ti,ab | 45,936 |
| 48 | geriatric:ti,ab | 5,730 |
| 49 | MeSH descriptor: [Aged] explode all trees | 213,642 |
| 50 | aged:ti,ab | 118,657 |
| 51 | frail:ti,ab | 2,397 |
| 52 | frails:ti,ab | 4 |
| 53 | frailty:ti,ab | 2,066 |
| 54 | MeSH descriptor: [Frailty] explode all trees | 198 |
| 55 | frailness:ti,ab | 0 |
| 56 | ageing:ti,ab | 9,339 |
| 57 | senior:ti,ab | 3,235 |
| 58 | elders:ti,ab | 1,199 |
| 59 | #46 OR #47 OR #48 OR #49 OR #50 OR #51 OR #52 OR #53 OR #54 OR #55 OR #56 OR #57 OR #58 | 381,978 |
| 60 | "hospital associated":ti,ab | 99 |
| 61 | "hospital-associated":ti,ab | 99 |
| 62 | "hospital acquired":ti,ab | 823 |
| 63 | "hospital-acquired":ti,ab | 823 |
| 64 | "hospital related":ti,ab | 60 |
| 65 | "hospital-related":ti,ab | 60 |
| 66 | "hospitalization associated":ti,ab | 82 |
| 67 | "hospitalization-associated":ti,ab | 82 |
| 68 | "hospitalisation-associated":ti,ab | 82 |
| 69 | "hospitalisation associated":ti,ab | 82 |
| 70 | "hospitalization acquired":ti,ab | 1 |
| 71 | "hospitalization-acquired":ti,ab | 1 |
| 72 | "hospitalisation acquired":ti,ab | 1 |
| 73 | "hospitalisation-acquired":ti,ab | 1 |
| 74 | "hospitalization related":ti,ab | 58 |
| 75 | "hospitalization-related":ti,ab | 58 |
| 76 | "hospitalisation related":ti,ab | 58 |
| 77 | "hospitalisation-related":ti,ab | 58 |
| 78 | "before hospitalisation":ti,ab | 128 |
| 79 | "before hospitalization":ti,ab | 128 |
| 80 | "after hospitalisation":ti,ab | 785 |
| 81 | "after hospitalization":ti,ab | 785 |
| 82 | functional deterioration near/3 hospital*:ti,ab | 16 |
| 83 | hospital* near/3 functional loss:ti,ab | 93 |
| 84 | functional status near/3 hospital*:ti,ab | 134 |
| 85 | los* near/3 independence:ti,ab | 250 |
| 86 | exposure near/2 hospital*:ti,ab | 74 |
| 87 | functional decline near/4 hospital*:ti,ab | 62 |
| 88 | #60 OR #61 OR #62 OR #63 OR #64 OR #65 OR #66 OR #67 OR #68 OR #69 OR #70 OR #71 OR #72 OR #73 OR #74 OR #75 OR #76 OR #77 OR #78 OR #79 OR #80 OR #81 OR #82 OR #83 OR #84 OR #85 OR #86 OR #87 | 2,574 |
| 89 | #10 AND #23 AND #45 AND #59 AND #88 | 222 |

### Appendix 2. Study flow diagram

**PRISMA 2020 flow diagram for new systematic reviews which included searches of databases and registers only**

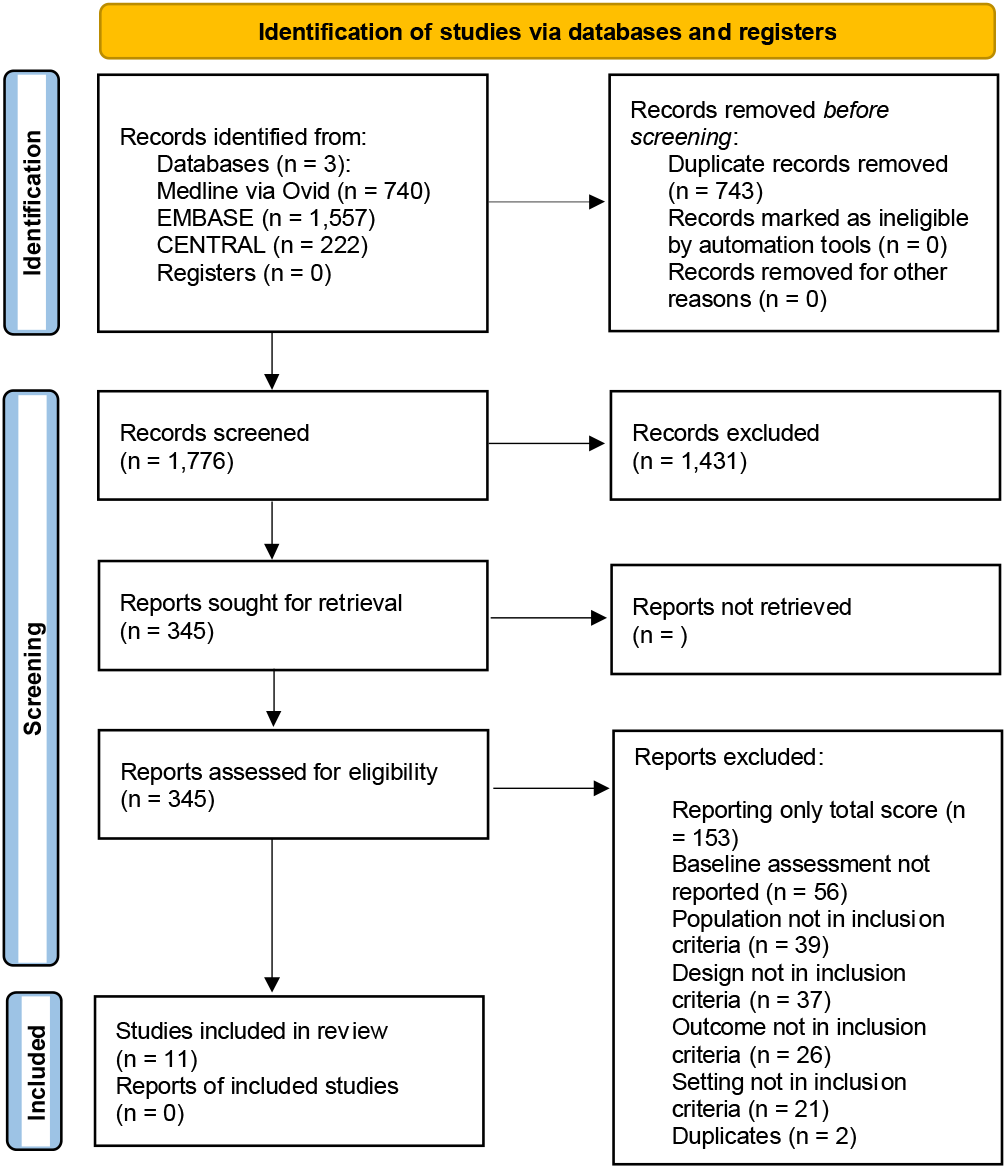

### Appendix 3. Individual question rating JBI Checklist

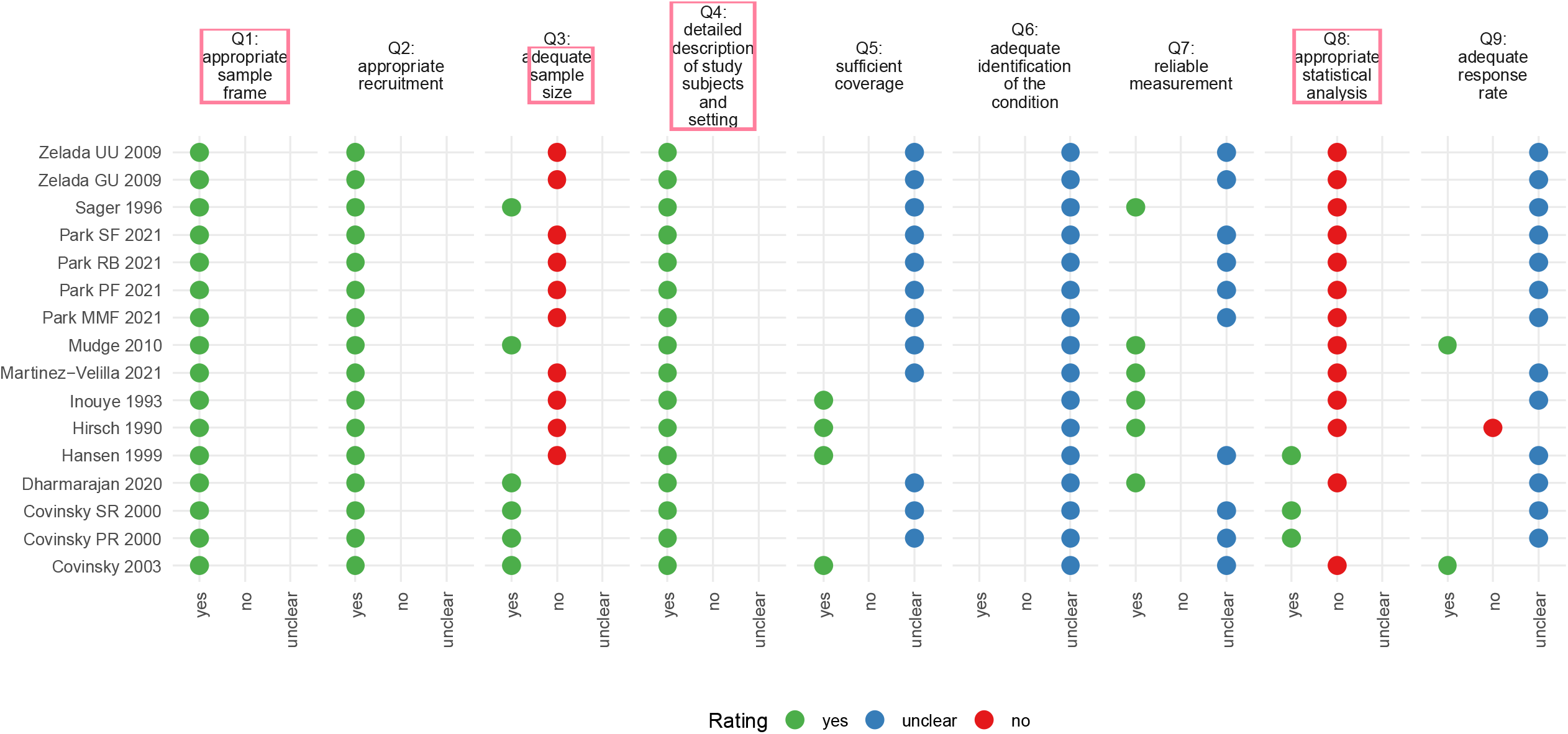

